# Relationship between sleep and progression of Parkinson’s disease – A Mendelian randomization study

**DOI:** 10.1101/2024.01.22.24301618

**Authors:** Mahiar Mahjoub, Elie Matar

## Abstract

**Background:** Sleep disturbances are common in Parkinson’s disease (PD) and growing evidence suggests a bidirectional relationship between sleep disruption and neurodegeneration.

**Objectives:** To study the causal relationship between sleep and rate of PD progression using two-sample Mendelian randomisation (MR).

**Methods:** Genetic variants linked to sleep duration and insomnia were analysed within a GWAS combining 12 longitudinal cohorts of patients with PD(n=4093 patients) examining motor and cognitive progression.

**Results:** Genetic liability to insomnia was associated with greater cognitive decline measured by MMSE. Consistent trends across MR estimates suggested a protective effect of increased sleep duration, and detrimental effect of insomnia on motor decline measured using UPDRS-III. Sensitivity analyses reinforced these relationships. The strength of causality among these associations was limited by heterogeneity and balanced pleiotropy.

**Conclusion:** Sleep related variables may alter the trajectory of cognitive and motor progression in PD and warrants further study.

## Introduction

Sleep disturbances are among the most common of the troubling non-motor symptoms of Parkinson’s disease (PD).^1–3^ Recently, there is growing recognition of the bidirectional influence of sleep disturbance on neurodegeneration, especially since the identification of the ‘glymphatic system’ through which sleep may facilitate the clearance of pathological proteins.^4,5^ Therefore, in addition to being a symptom of the disorder, it has been proposed that sleep disturbance may itself influence the natural trajectory of PD, and therefore represent a modifiable risk factor for disease progression^1^. However, dissecting out the causal influence of sleep disturbance on PD progression is complex, especially as sleep disturbances can occur early in PD and precede its diagnosis by several decades.^6^ Furthermore, practical, and ethical challenges limit the ability to implement large scale randomized placebo-controlled trials assessing the impact of sleep disturbance on a slowly progressing illness such as PD.^7^

Mendelian randomization (MR) is a powerful technique that can be used to infer causality between variables that may be intractable to traditional experimental designs.^8–10^ MR relies on the natural randomisation (since birth) of participants by alleles that have been linked to an exposure variable of interest.^7^ The proliferation of open-source genome-wide association studies (GWAS) has enabled the use of MR in the investigation of causation between many variables of interest including between sleep disturbances and neurodegenerative conditions such as PD.^9,11^ Recent MR studies have found negative or mixed results linking sleep and risk of developing PD and age of diagnosis respectively.^12^ However, whether a causal relationship exists between sleep variables and motor and cognitive progression of PD is unknown. In this study, we used two-sample MR to investigate the causal role between sleep and PD progression. Given the importance of sleep in the clearance of toxic and pathological extracellular proteins in other neurodegenerative models^4^, we hypothesized that genetic predisposition towards longer sleep would be protective and associated with slower rate of PD progression, whilst the converse would be expected in those with a predisposition towards reduced sleep (insomnia).

## Methods

### Exposure dataset

Exposure variables of interest including insomnia (N=462,341; ukb-b-3957) and sleep duration (N=460,099; ukb-b-4424) were obtained from the IEU OpenGWAS database based on UK BioBank data.^13^ Both variables were self-reported. Instrumental Variables (IVs) were selected based on SNVs with p-value <5×10^−8^. Variants lacking rsID formatting were removed to avoid mismatching.

### Outcome dataset

Outcome variables were derived from a meta-GWAS analysis of 12 longitudinal cohorts of patients with PD (N=4093, 22307 observations, median follow-up=3.8 years).^14^ Markers of cognitive progression included change in Montreal Cognitive Assessment (MOCA) and Mini-Mental State Examination (MMSE) scores. Motor progression was based on the Unified Parkinson’s Disease Rating Scale Part III (UPDRS-III).

### Harmonisation

Proxy SNVs were used to increase the search space of matching IVs between the exposure and outcome variables (R^2^>0.8 and 10000 base pairs). The European 1000 Genomes Linkage Disequilibrium (LD) reference panel was used for the proxy search.^15^ The R package *gwasvcf* was used for proxy search.^16^ Palindromic IVs were removed. Clumping (clumping window = 10000 base pairs, R2 <0.001) was performed to remove IVs in LD. Clumping was done at this stage as opposed to the IV selection stage to increase the chance of finding matching IVs.^12^

### Statistical analysis

Two-sample MR was performed using the Inverse-Variant-Weighted (IVW) method to derive the effect estimate.^9,17,18^ A causative relationship between an exposure and an outcome variable was defined as a statistically significant beta effect estimate (two-sided p-value < 0.05) obtained by the IVW method. All analysis was performed in the R language environment with the *TwoSampleMR* package used for MR analysis.^7,19^ The PhenoScanner variant database was used to query variant traits.^20,21^ Sensitivity analyses was performed. More information on the sensitivity analysis and inference can be found in the Supplementary Methods.

### Data availability

The study can be reproduced by using the cited R packages. All data used is open-source.

## Results

### Sleep and cognitive progression in PD

Effect estimates across MR methods demonstrated a consistent relationship suggesting insomnia variants were associated with a greater rate of cognitive decline using MMSE. IVW effect estimates were statistically significant as were estimates derived from the outlier robust methods, such as Weighted-Median (WM), MR-PRESSO (MRP), contamination mixture (CM) and lasso methods (Table 1, Figures S2, S6). However, the non-significant Egger result suggested the presence of balanced pleiotropy (Table 1, Figure S2). There was no directional pleiotropy and minimal heterogeneity (Q-Cochrane score=2.3, p-value=0.81) (Figures S4-S6). There was no clear relationship between sleep duration and MMSE across all methods (Table 1, Figures S3, S10).

**Table 1.** Effect estimates of sleep variables and PD progression markers (95% CI).

| Relationship | IVW | WM | MRPc | Egger |
| --- | --- | --- | --- | --- |
| **Insomnia vs.** |  |  |  |  |
| UPDRS-III | 2.4 (-1.8,6.6) | **5.8 (2.8,8.8) \*** | **5.1 (1.3,8.9) \*** | -4.7 (-39.2,29.8) |
| UPDRS-III<sup>+</sup> | **4.0 (4x10<sup>-4</sup>,8.1) \*** | **6.0 (3.1,9.0) \*** | **5.6 (2.3,8.9) \*** | -1.7 (-32.7,29.2) |
| MMSE | **13.3 (8.7,17.8)\*** | **13.4 (7.5,19.3)\*** | NA<sup>#</sup> | 6.8 (-43.7,57.4) |
| MOCA | -4.6 (-43.4,34.1) | -16.1 (-38.2,6.1) | NA<sup>#</sup> | -26.1 (-137.1,85.0) |
| **Sleep duration vs.** |  |  |  |  |
| UPDRS-III | -3.3 (-7.4,0.9) | **-5.4 (-8.7,-2.2) \*** | **-5.0 (-8.9,-1.1) \*** | -0.1 (-29.8,29.6) |
| UPDRS-III<sup>^</sup> | **-5.0 (-8.9,-1.1) \*** | **-5.8 (-9.1,-2.6) \*** | **-7.0 (-9.0,-4.9) \*** | -4.5 (-30.5,21.5) |
| MOCA | -14.5 (-32.4,3.3) | **-24.4 (-36.4,-12.4) \*** | **-27.6 (-34.6,-20.6) \*** | -10.3 (-105.7,85.2) |
| MOCA<sup>a</sup> | **-21.5 (-37.1,-5.9) \*** | **-24.7 (-36.7,-12.7) \*** | **-27.6 (-34.6,-20.6) \*** | -39.6 (-120.5,41.4) |
| MOCA<sup>b</sup> | **-18.9 (-35.2,-2.6) \*** | **-24.6 (-36.7,-12.4) \*** | **-27.6 (-34.6,-20.6) \*** | 31.1 (-51.9,114.1) |
| MOCA<sup>c</sup> | **-27.6 (-37.4,-17.8) \*** | **-24.9 (-37.6,-12.2) \*** | NA<sup>#</sup> | 1.8 (-47.5,51.2) |
| MMSE | -0.2 (-7.3,6.9) | 0.2 (-5.2,5.7) | 3.5 (-4.0,10.9) | -35.2 (-89.3,18.8) |

Abbreviations: MOCA=Montreal Cognitive Assessment; MMSE=Mini-Mental State Examination; UPDRS-III=Unified Parkinson's Disease Rating Scale Part III; IVW=inverse-variant weighted; WM=weighted median; MRPc=MR-PRESSO.
<sup>+</sup> outlier variant rs4379719 excluded
<sup>^</sup> outlier variant rs1633106 excluded
<sup>a</sup> outlier variant rs16959955 excluded
<sup>b</sup> outlier variant rs2236295 excluded
<sup>c</sup> both outliers rs16959955 and rs2236295 excluded
\*P-value<0.05
<sup>#</sup> unable to achieve minimum threshold

Non-significant IVW effect estimates were obtained when examining the relationship between variables of sleep and MOCA (Table 1, Figures S2-S3). In the case of insomnia, this was supported by WM and Egger methods with significant heterogeneity amongst the four IVs used (Figures S2-S4, S7). However, a consistent inverse relationship between sleep duration and cognitive decline (measured using MOCA), was seen across sensitivity analyses except using Egger’s method (Table 1, Figure S3) and significant heterogeneity (Q-Cochrane score=30.9, p-value=6.6×10^−5^) and balanced pleiotropy was present (Figures S4-S5). LOO analysis identified rs16959955 and/or rs2236295 as potential pleiotropic variants (Figure S13). Their removal led to a significant effect estimate by IVW and all sensitivity analysis methods except Egger (Table 1, Figures S3,S14-S16). Removal of variants eliminated significant heterogeneity however balanced pleiotropy remained present (Figures S4-S6). A database search of the phenotypes associated with the two variants did not reveal a clear mechanism underpinning the pleiotropic effects (see supplementary materials).

### Sleep and motor progression in PD

The directionality across the different MR estimates suggested a consistent protective effect of increased sleep duration, and detrimental effect of insomnia on motor decline in PD. Insomnia variants correlated with more rapid decline in UPDRS-III scores over time, whilst this directionality was reversed for variants associated with increasing sleep duration (Table 1, Figures S2-S3). A trend was seen in the initial IVW estimates, which did not cross statistical significance. However, sensitivity analysis suggested significant heterogeneity (Q-Cochrane score >30, p-value < 0.05) with balanced pleiotropy (Figures S4-S5). Outlier robust estimates (WM, MRP, CM and lasso methods) all demonstrated consistent statistical significance, as did the IVW estimates after LOO analysis (Table 1, Figures S2-S3). Specifically, removal of the variant rs1633106 resulted in a statistically significant IVW effect estimate between sleep duration and reduced rate of motor progression (Table 1, Figures S12). Similarly, removal of rs4379719 resulted in a significant effect estimate by IVW between insomnia and increasing motor decline (Table 1, Figure S9). Variant database search showed that rs4379719 was also correlated with increasing naps suggesting a clear source of pleiotropic effect. However, significant heterogeneity and possible balanced pleiotropy remained (Figures S4-S5). The Egger estimate remained non-significant across comparisons as would be expected in the presence of heterogeneity and pleiotropy, even if balanced (Table 1, Figure S5).

## Discussion

This study used two-sample MR to uniquely probe the causal influence of sleep and its disturbance on cognitive and motor decline in PD. The directionality of our findings is in keeping with our hypotheses. The predominant finding based on significant initial MR estimates suggested that a causal role between the presence of insomnia and cognitive decline (change in MMSE) is likely present. Likewise, insomnia was associated with increased motor progression (change in UPDRS-III score) across a number of MR estimates, although statistical significance in the IVW estimate was reached only after the removal of pleiotropic variants.

The relationship between sleep duration and cognitive decline was less robust, although the directionality was consistent with a potentially protective effect of sleep duration and cognitive decline (change in MOCA). Statistical significance was reached in IVW only after sensitivity analyses. Increasing sleep duration, however, was associated also with reduced rate of motor decline across several MR estimates, with significance reached again after removal of pleiotropic variants.

It is important to note that the certainty of the above causal relationships, supported by most MR methods in the analyses above is attenuated by the non-significant Egger estimates across comparisons. The Egger method is sensitive to the presence of pleiotropy and has lower power especially in the presence of heterogeneity.^22^ In all cases, this pleiotropy was found to be balanced and non-directional (thus not breaking MR assumptions). Sensitivity analysis revealed that heterogeneity and pleiotropy were driven especially by specific variants. An example of a known pleiotropic effect was seen in the case of the insomnia variant rs4379719, which has been associated with a napping phenotype (effect estimate = -0.007432, p-value=4.34×10^−7^). Naps are thought to contribute to insomnia due to reduced sleep pressure and can offset any sleep lost during the night. Accordingly, we expect this should mitigate the deleterious effects of insomnia on neurodegeneration, which was consistent with the effect of this variant on nullifying the MR outcome.

The above highlights an inherent limitation of the approach of this study. It is challenging to limit heterogeneity when using public GWAS databases for IV selection due to differences in the populations and methods used across the individual studies to derive the associations between genetic variants and risk factors. Pleiotropic effects due to other, as yet undiscovered biological functions and phenotypes of the variants used in the study is also possible. Furthermore, available GWAS data on sleep phenotypes have typically used subjective questionnaire data, which may not accurately reflect the true amount of sleep of an individual.^23^ Using GWAS data linked to more objective instruments such as actigraphy may strengthen the associations in this study, as will the use of GWAS studies with larger sample sizes. Discovery and cataloguing of more variants over time will increase the inclusion of valid IVs and hopefully reduce the effects of heterogeneity.

Our findings are encouraging and support the growing body of evidence suggesting an active influence of sleep on neurodegeneration, which is speculated to occur through a number of mechanisms such as impaired glymphatic clearance of pathological proteins, altered protein homeostasis, and inflammation.^1^ This raises the possibility that identifying and treating sleep disturbance may modify the disease trajectory of PD. Significant heterogeneity and pleiotropic effects rendered some causal links uncertain and warrant future MR studies targeting sleep in neurodegeneration.

## Authors’ Roles

MM – Execution, analysis, writing. EM – Design, execution, editing of the final version of the manuscript

## Financial disclosures of all authors for the preceding 12 months

MM – MM is currently employed by the Sydney Local Health District, Government of New South Wales.

EM – EM was funded by an NHMRC Grant #2008565. EM has received honoraria from the International Movement Disorders Society (MDS) and CSL Seqirus. EM is currently employed by the University of Sydney.

