## Supplementary Methods for "Relationship between sleep and progression of Parkinson’s disease – A Mendelian randomization study"

Supplementary Material

### Supplementary Methods

*Statistical analysis*

Two-sample MR was performed using the Inverse-Variant-Weighted (IVW) method to derive the effect.^1–3^ A causative relationship between an exposure and an outcome variable was defined as a statistically significant beta effect estimate (two-sided p-value < 0.05) obtained by the IVW method.

*Sensitivity analysis*

Sensitivity analysis was performed using consensus and outlier robust methods.^3–5^ Consensus methods included weighted median (WM) and Egger. Outlier robust methods included MR-PRESSO (MRP), MR-LASSO (lasso) and consensus mixture (CM). Heterogeneity was assessed using Q-Cochrane score.^3^ The Egger y-intercept was used to probe directional pleiotropy.^6^ Scatter plots and LOO analysis were used to help assess heterogeneity and pleiotropy.^3,7^ The PhenoScanner variant database was used to query variants suspected of involvement in pleiotropy based on the LOO analysis.^3^ A summary of the statistical inference framework used in this study is shown in Figure S1.

*Inference*

Each method used as part of the sensitivity analysis accounted for various assumptions of MR analysis and IV selection.^3^ A statistically significant effect estimate based on the WM method ensured the eligibility of the majority of IVs (>50%).^5^ The presence of horizontal pleiotropy was suggested by a statistically significant non-zero Egger y-intercept.^4^ Heterogeneity was defined as a statistically significant Q-cochrane score. Increasing heterogeneity increased the likelihood of potentially pleiotropic variants.^3,4^ A statistically non-significant non-zero Egger y-intercept despite the presence of significant heterogeneity suggested bidirectional (or balanced) pleiotropy unlikely to violate MR assumptions.^4,8^ Leave-one-out (LOO) analysis was performed to ensure that a causal relationship was not dominated by any one IV.^3^ It was also performed in cases where statistically non-significant estimates were obtained by IVW, and/or in other methods, to help in the identification of pleiotropic variants.

### Figure S1: Pipeline used for MR analysis and inference.

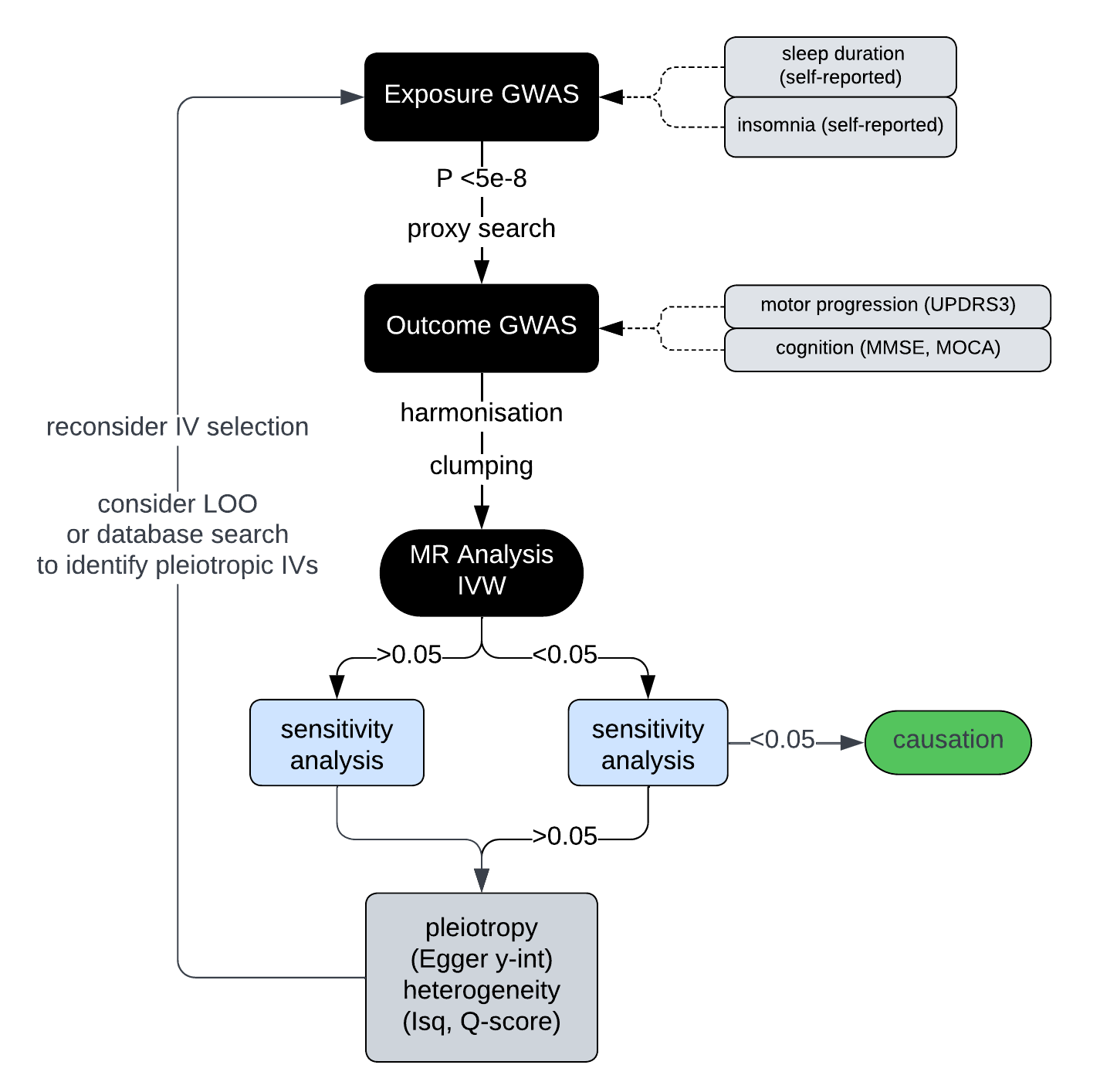

Abbreviations: MMSE=Mini Mental State Examination; MOCA=Montreal Cognitive Assessment; UPDRS= Unified Parkinson's Disease Rating Scale; IVW=inverse-variance weighted; LOO=leave-one-out analysis; IV=instrumental variable

### Figure S2: Forest plot of insomnia (self-reported) and PD progression markers.

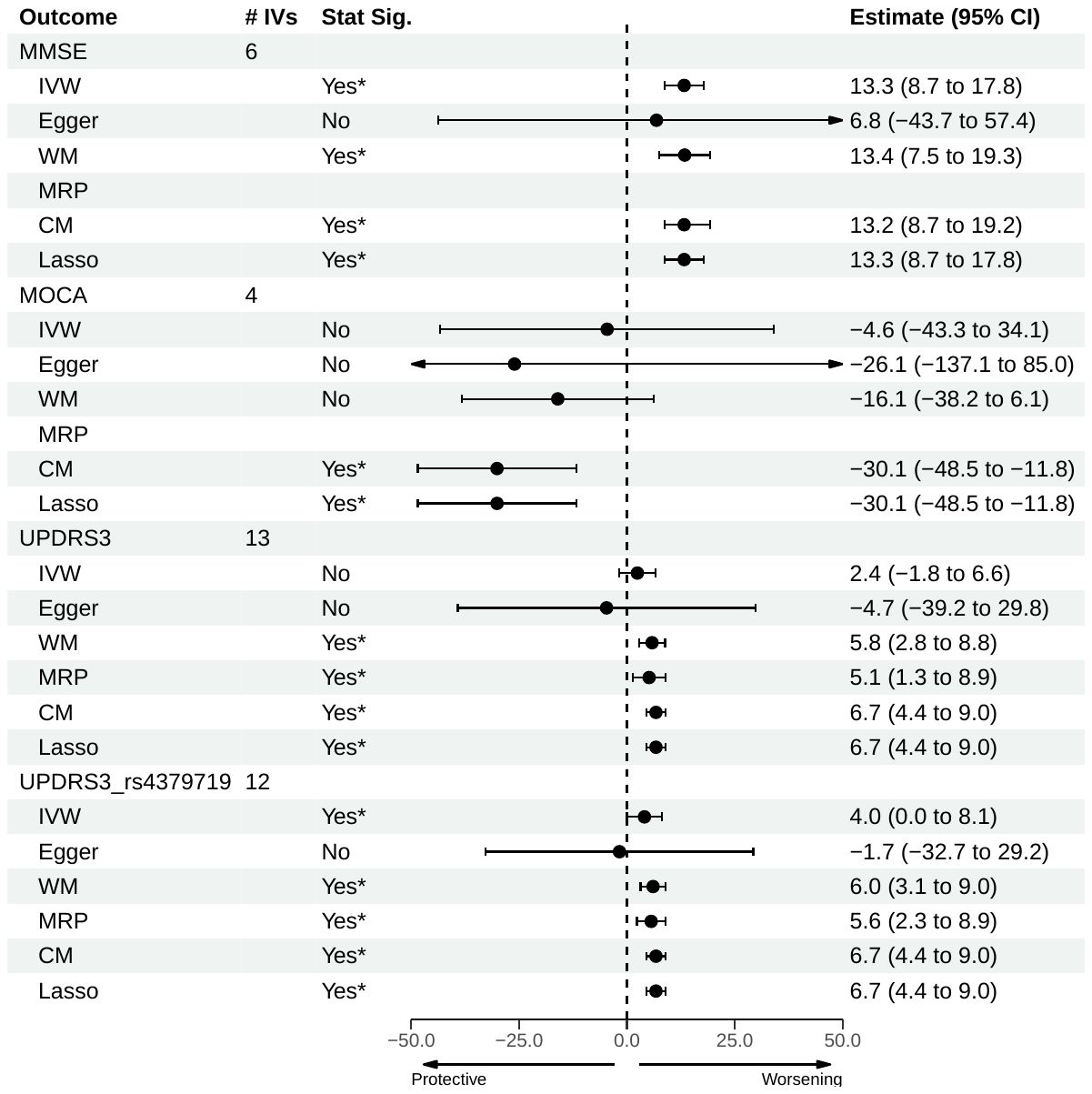

Abbreviations: MOCA=Montreal Cognitive Assessment; MMSE=Mini-Mental State Examination; UPDRS-III=Unified Parkinson's Disease Rating Scale Part III; IVW=inverse-variance weighted; WM=weighted median; MRP=MR-PRESSO; CM=contamination mixture; CI=confidence interval

The confidence intervals correspond to beta estimates obtained from each corresponding method of MR analysis.

UPDRS3_rs4379719 refers to MR study after removal of rs4379719.

### Figure S3: Forest plot of sleep duration (self-reported) and PD progression markers.

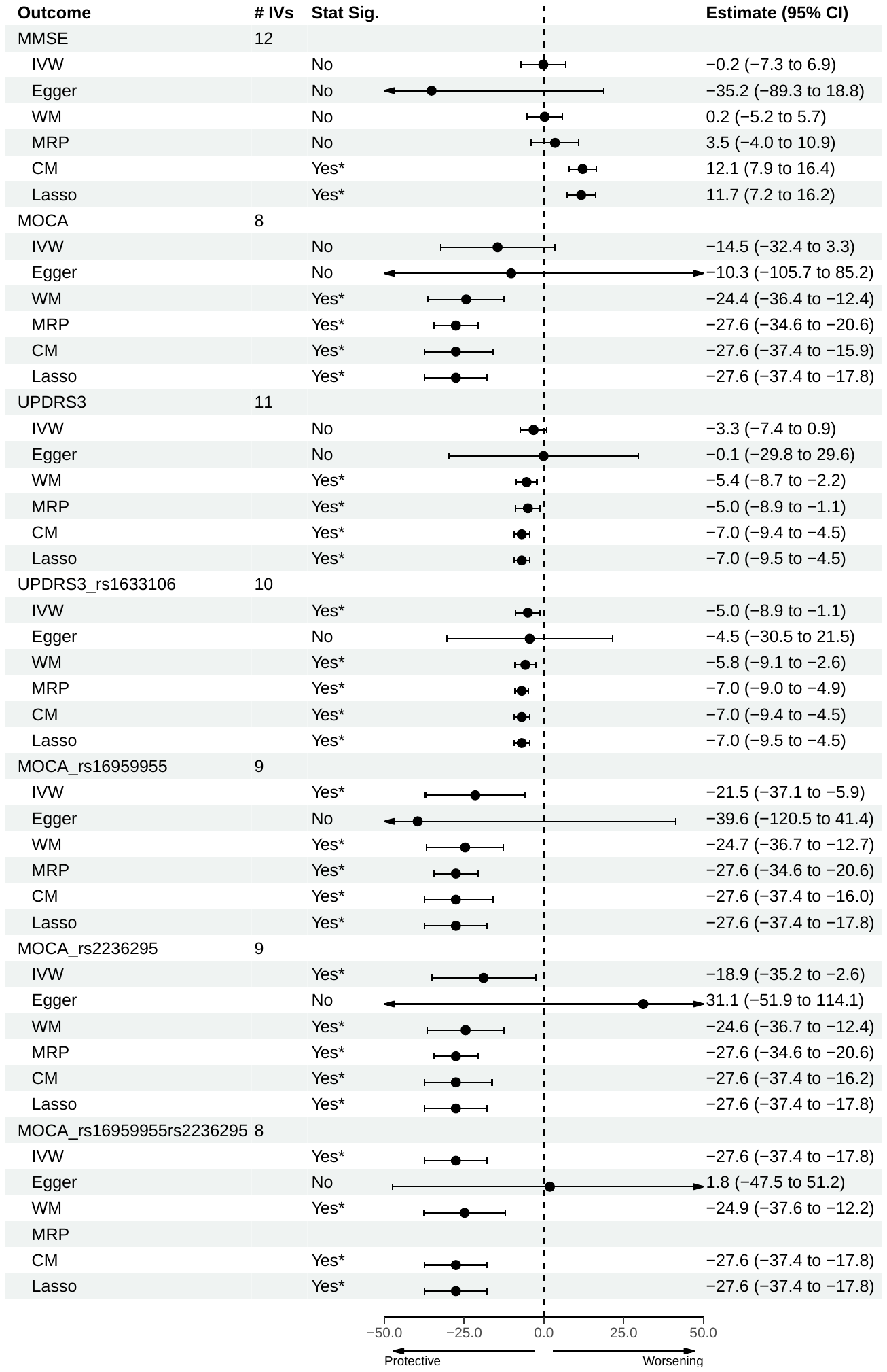

Abbreviations: MOCA=Montreal Cognitive Assessment; MMSE=Mini-Mental State Examination; UPDRS-III=Unified Parkinson's Disease Rating Scale Part III; IVW=inverse-variance weighted; WM=weighted median; MRP=MR-PRESSO; CM=contamination mixture; CI=confidence interval

The confidence intervals correspond to beta estimates obtained from each corresponding method of MR analysis.

Where an IV is included in the outcome label, the corresponding IV was removed from the study as it was identified as potentially pleiotropic based on sensitivity and leave-one-out analysis.

### Figure S4: Q-Cochrane scores for each MR analysis to assess heterogeneity, calculated by both IVW and Egger methods.

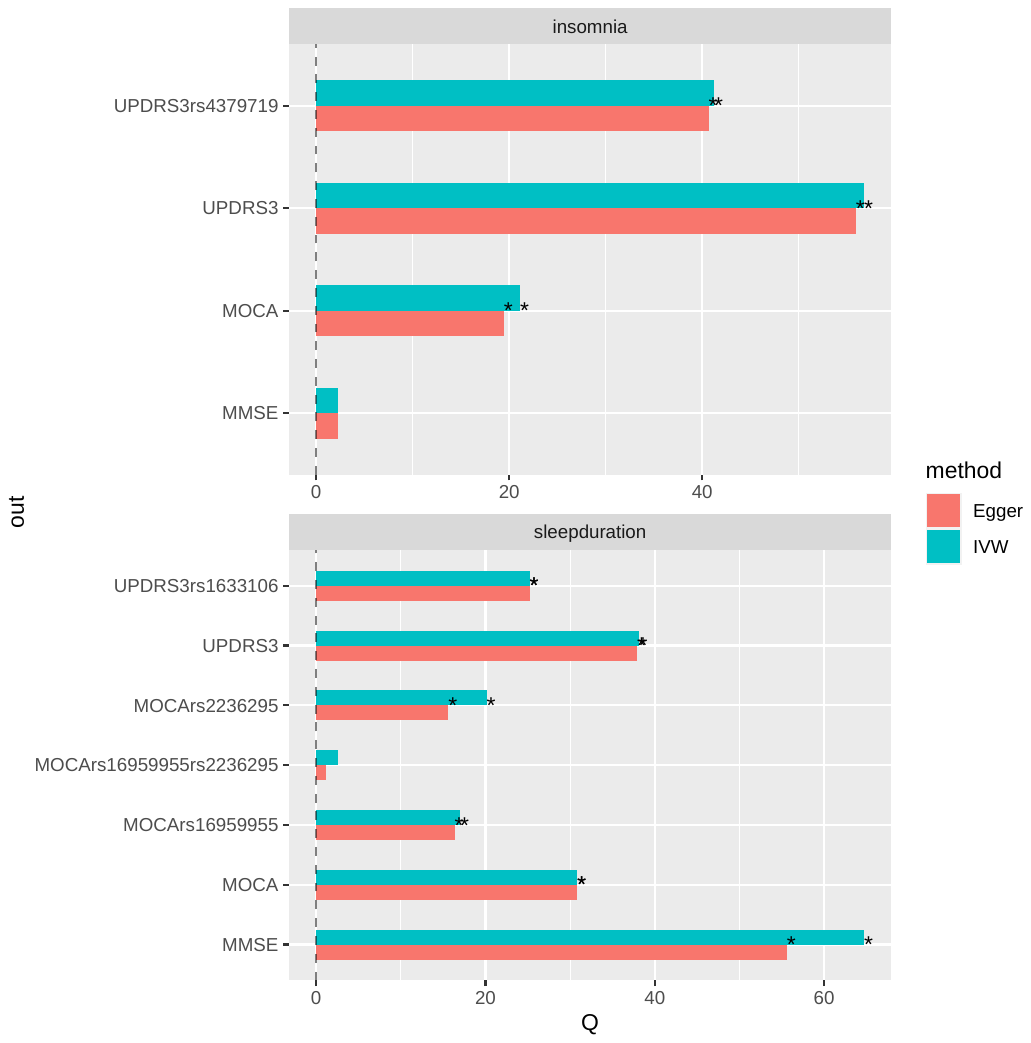

Abbreviations: Q=Q-cochrane score; out=outcome variable; IVW=inverse-variance weighted; MOCA=Montreal Cognitive Assessment; MMSE=Mini-Mental State Examination; UPDRS-III=Unified Parkinson's Disease Rating Scale Part III

*statistical significance for heterogeneity based on Q-Cochrane score (<0.05).

### Figure S5: Egger y-intercept values obtained from the MR analysis to assess horizontal pleiotropy.

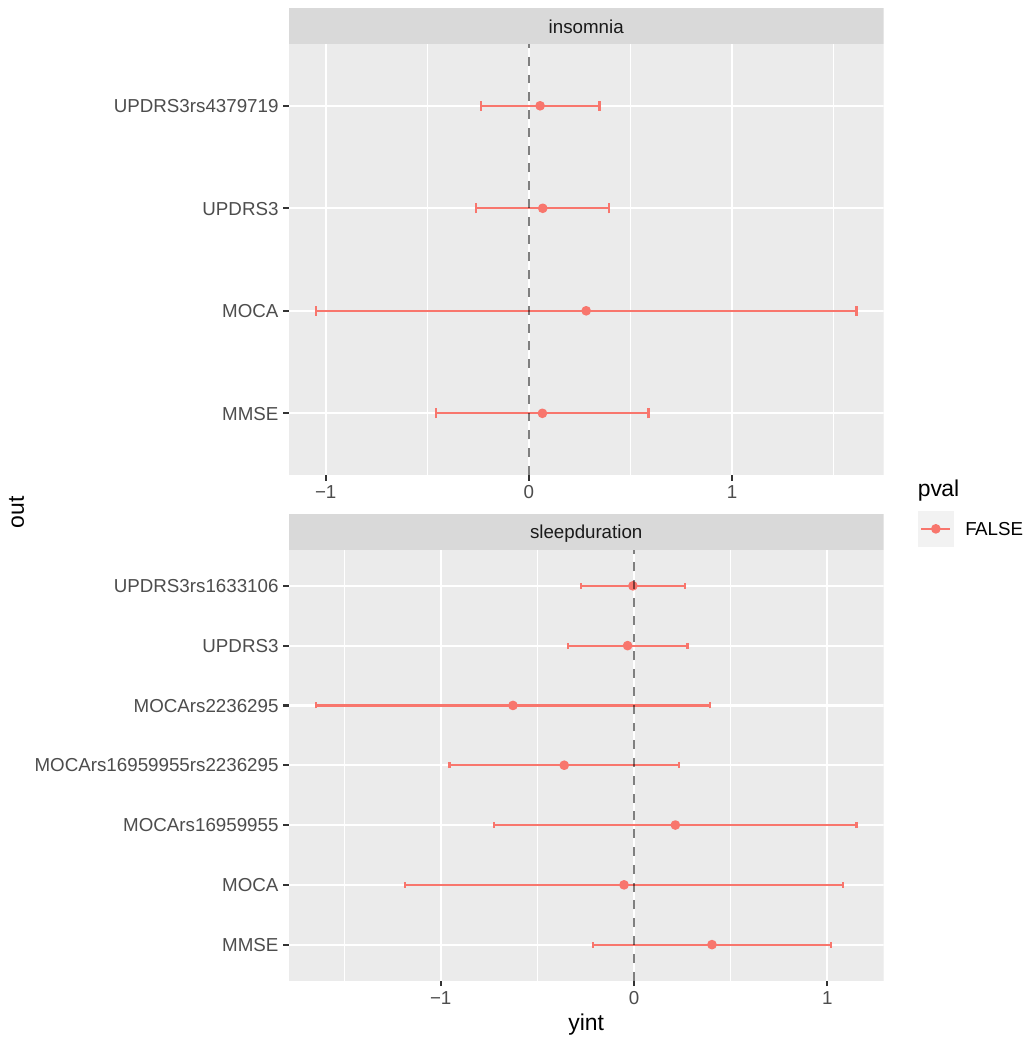

Abbreviations: yint=y-intercept (Egger); out=outcome variable; pval=p-value significance; MOCA=Montreal Cognitive Assessment; MMSE=Mini-Mental State Examination; UPDRS-III=Unified Parkinson's Disease Rating Scale Part III

The errors bars correspond to 95% confidence intervals.

*denotes statistical significance of a non-zero y-intercept (<0.05).

### Figure S6: Plots used to aid causal inference based on MR for insomnia vs MMSE

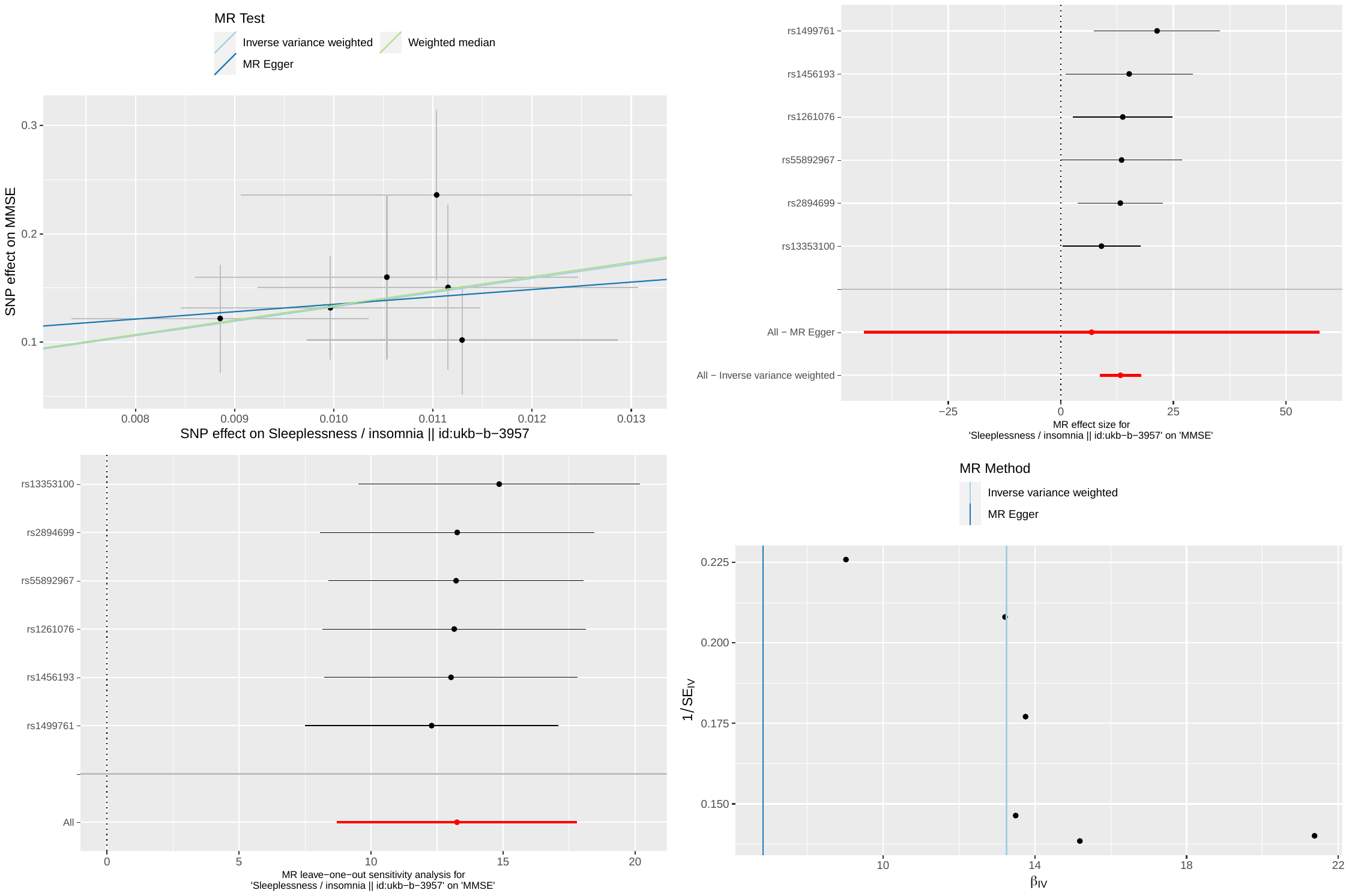

(top left) Scatter plot of the exposure (x-axis) and outcome (y-axis) variables of interest. Each point denotes a single IV included in the study. The x- and y-values of each point correspond to the effect estimate of each IV to the exposure and outcome variable, respectively. The gradient of each regression line corresponds to the effect estimate obtained by each method (IVW, Egger, WM).

(top right) Forest plots of the effect estimates of each single IV on the outcome. Pooled effect estimate based on Egger and IVW methods are shown by the red bars.

(bottom left) Leave-one-out analysis. Red bar denotes the effect estimate by IVW.

(bottom right) Funnel plot used to gauge directional pleiotropy suggested by asymmetry.

All error bars correspond to 95% confidence intervals.

### Figure S7: Plots used to aid causal inference based on MR for insomnia vs MOCA

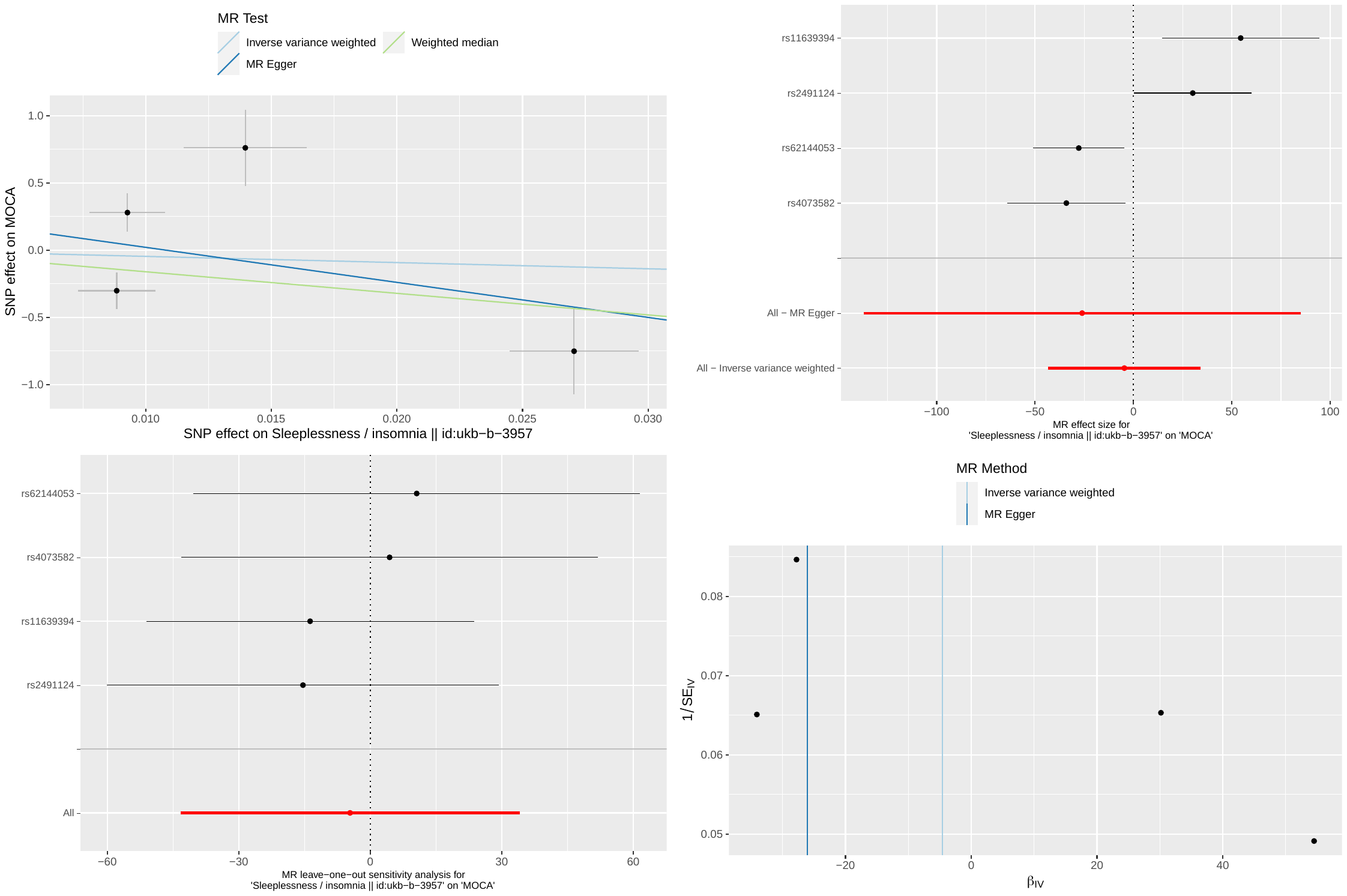

(top left) Scatter plot of the exposure (x-axis) and outcome (y-axis) variables of interest. Each point denotes a single IV included in the study. The x- and y-values of each point correspond to the effect estimate of each IV to the exposure and outcome variable, respectively. The gradient of each regression line corresponds to the effect estimate obtained by each method (IVW, Egger, WM).

(top right) Forest plots of the effect estimates of each single IV on the outcome. Pooled effect estimate based on Egger and IVW methods are shown by the red bars.

(bottom left) Leave-one-out analysis. Red bar denotes the effect estimate by IVW.

(bottom right) Funnel plot used to gauge directional pleiotropy suggested by asymmetry.

All error bars correspond to 95% confidence intervals.

### Figure S8: Plots used to aid causal inference based on MR for insomnia vs UPDRS3

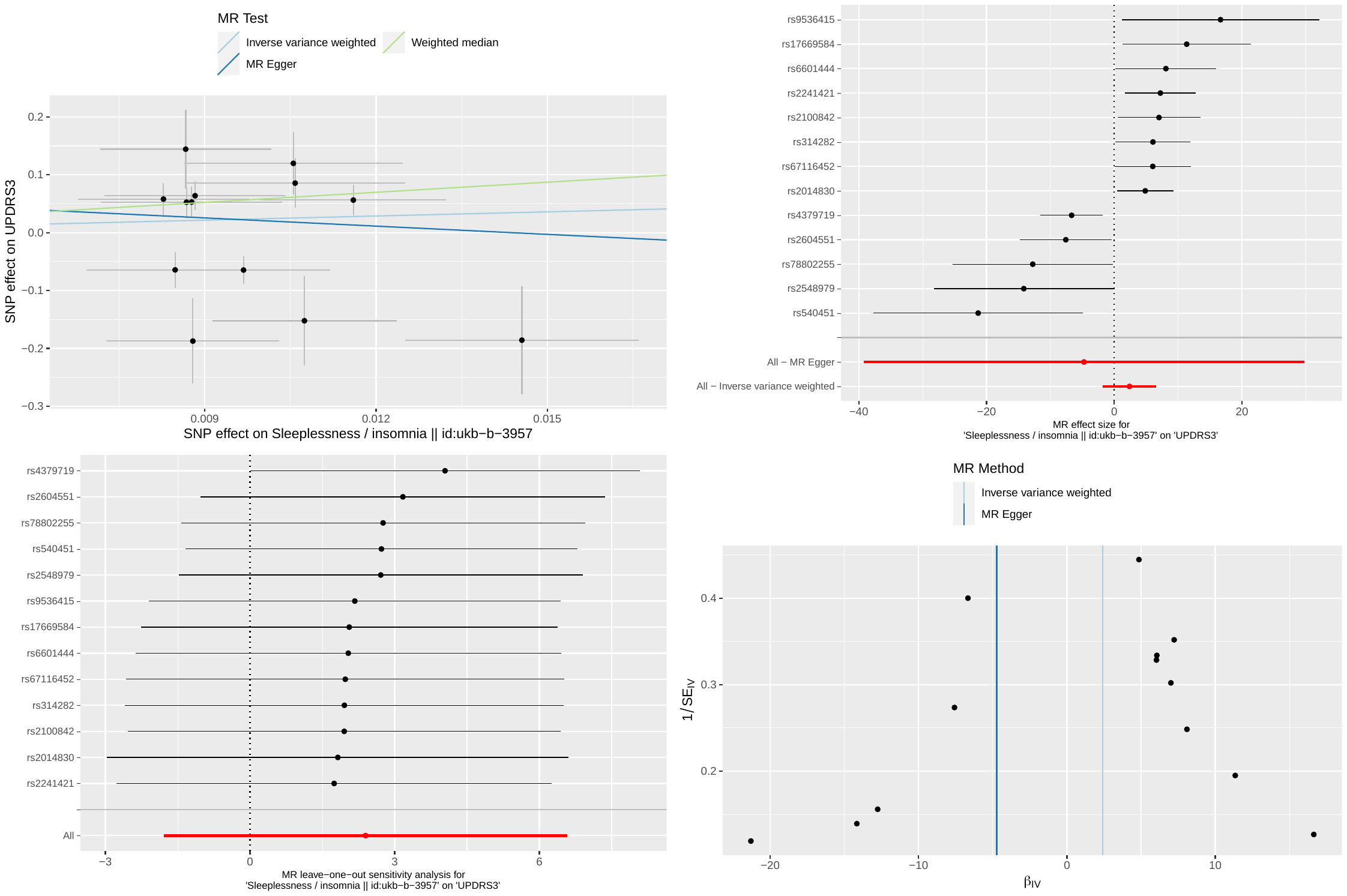

(top left) Scatter plot of the exposure (x-axis) and outcome (y-axis) variables of interest. Each point denotes a single IV included in the study. The x- and y-values of each point correspond to the effect estimate of each IV to the exposure and outcome variable, respectively. The gradient of each regression line corresponds to the effect estimate obtained by each method (IVW, Egger, WM).

(top right) Forest plots of the effect estimates of each single IV on the outcome. Pooled effect estimate based on Egger and IVW methods are shown by the red bars.

(bottom left) Leave-one-out analysis. Red bar denotes the effect estimate by IVW.

(bottom right) Funnel plot used to gauge directional pleiotropy suggested by asymmetry.

All error bars correspond to 95% confidence intervals.

### Figure S9: Plots used to aid causal inference based on MR for insomnia vs UPDRS3, without rs4379719

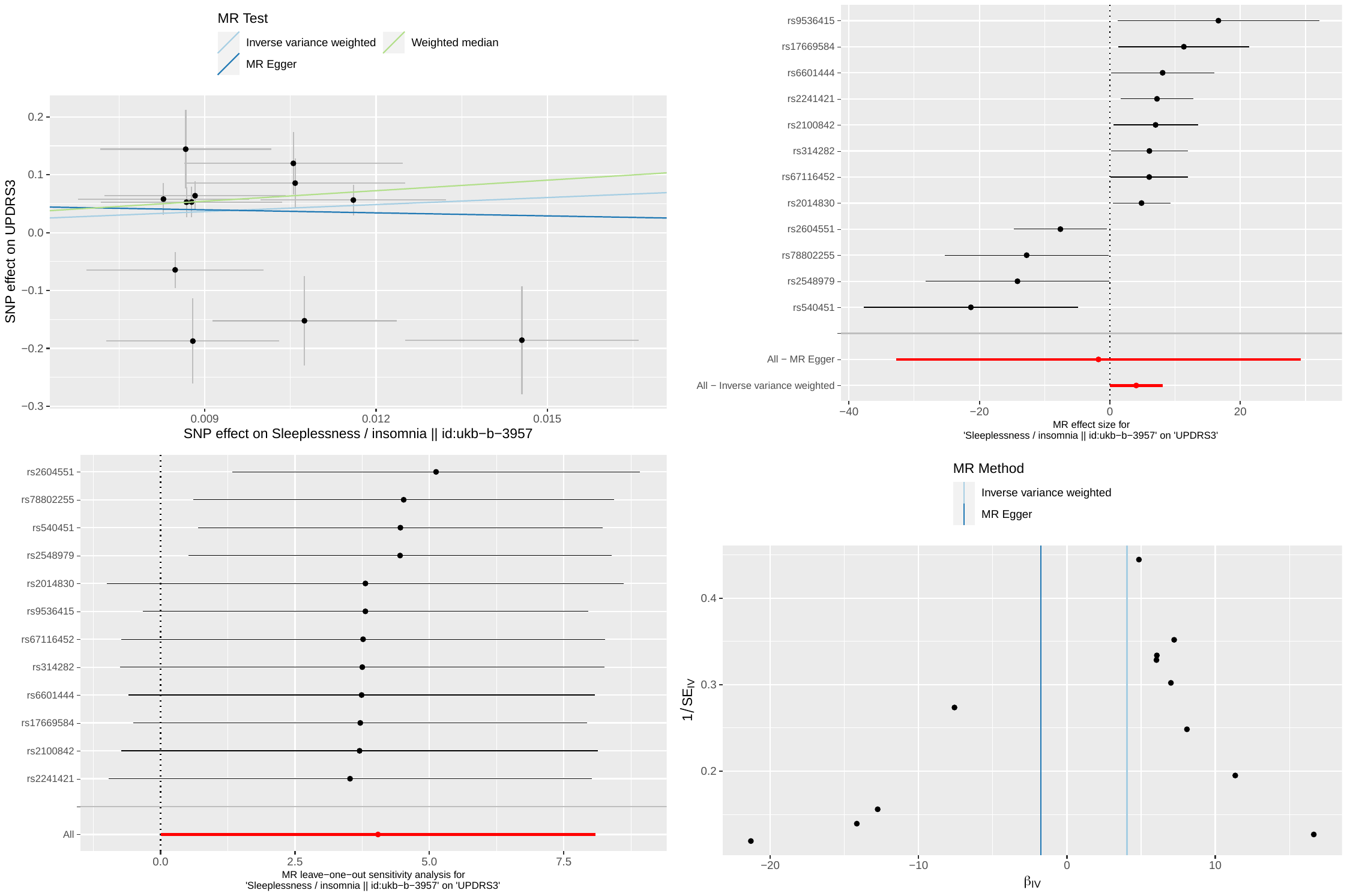

(top left) Scatter plot of the exposure (x-axis) and outcome (y-axis) variables of interest. Each point denotes a single IV included in the study. The x- and y-values of each point correspond to the effect estimate of each IV to the exposure and outcome variable, respectively. The gradient of each regression line corresponds to the effect estimate obtained by each method (IVW, Egger, WM).

(top right) Forest plots of the effect estimates of each single IV on the outcome. Pooled effect estimate based on Egger and IVW methods are shown by the red bars.

(bottom left) Leave-one-out analysis. Red bar denotes the effect estimate by IVW.

(bottom right) Funnel plot used to gauge directional pleiotropy suggested by asymmetry.

All error bars correspond to 95% confidence intervals.

### Figure S10: Plots used to aid causal inference based on MR for sleep duration vs MMSE

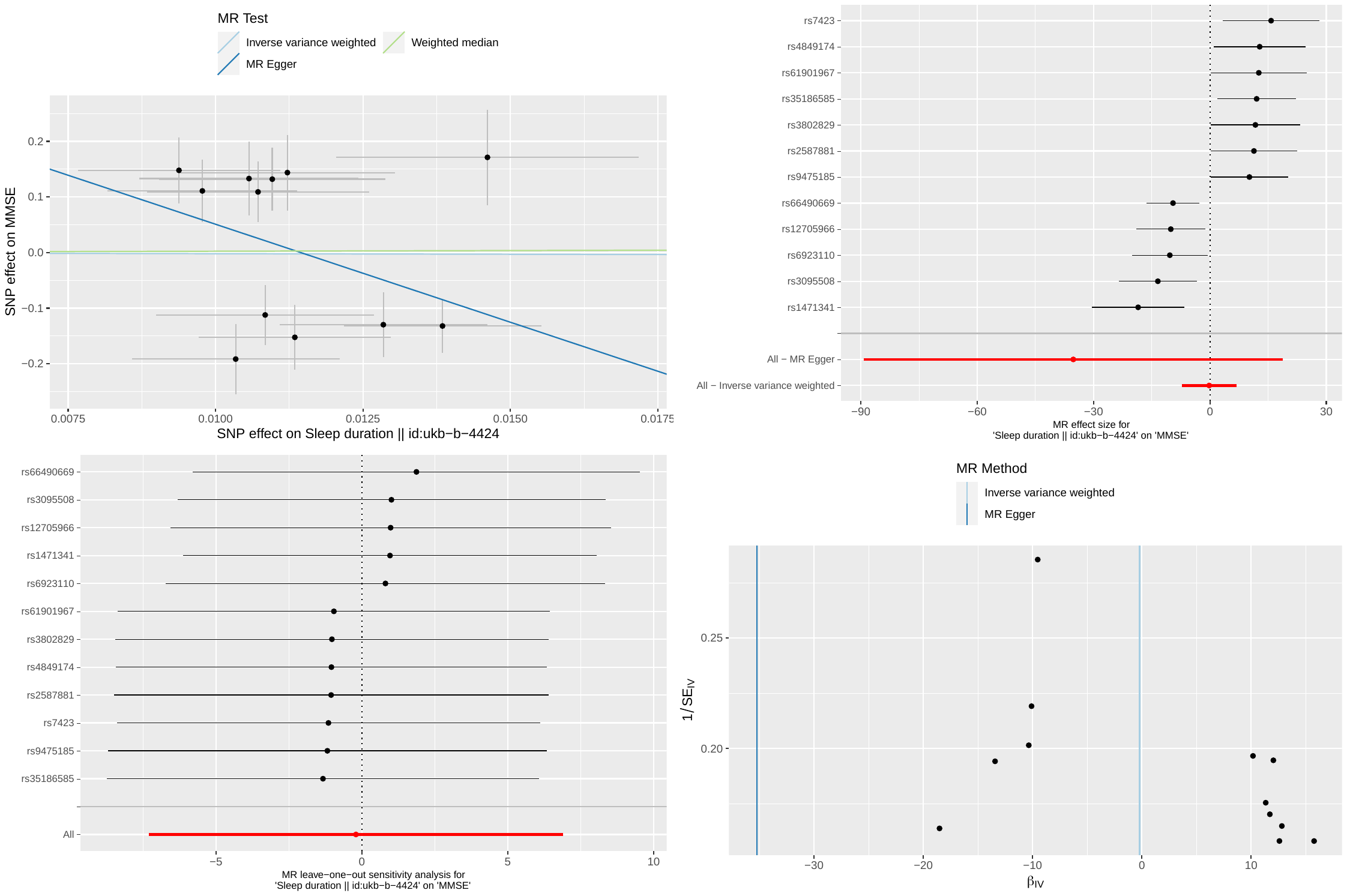

(top left) Scatter plot of the exposure (x-axis) and outcome (y-axis) variables of interest. Each point denotes a single IV included in the study. The x- and y-values of each point correspond to the effect estimate of each IV to the exposure and outcome variable, respectively. The gradient of each regression line corresponds to the effect estimate obtained by each method (IVW, Egger, WM).

(top right) Forest plots of the effect estimates of each single IV on the outcome. Pooled effect estimate based on Egger and IVW methods are shown by the red bars.

(bottom left) Leave-one-out analysis. Red bar denotes the effect estimate by IVW.

(bottom right) Funnel plot used to gauge directional pleiotropy suggested by asymmetry.

All error bars correspond to 95% confidence intervals.

### Figure S11: Plots used to aid causal inference based on MR for sleep duration vs UPDRS3

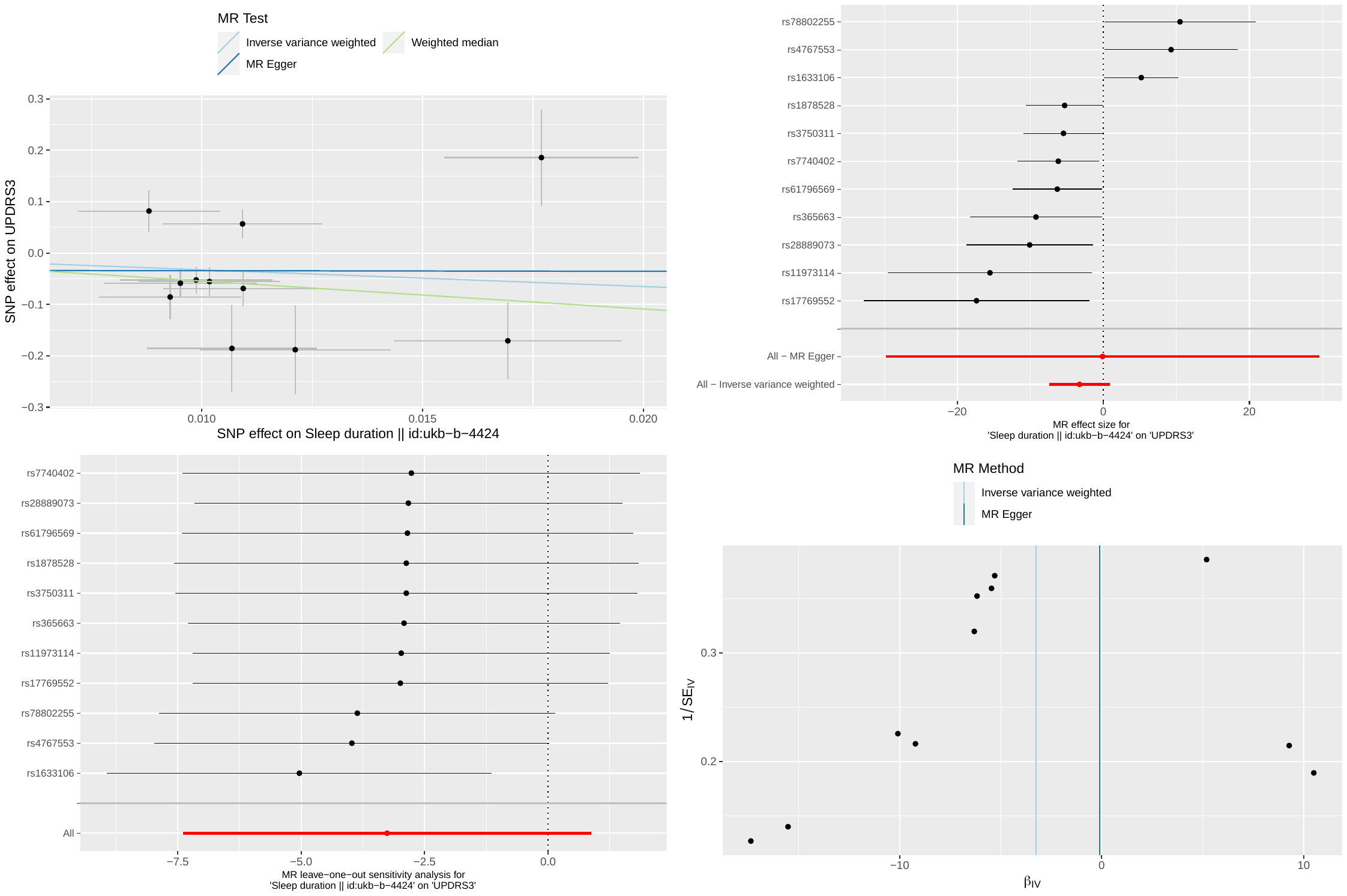

(top left) Scatter plot of the exposure (x-axis) and outcome (y-axis) variables of interest. Each point denotes a single IV included in the study. The x- and y-values of each point correspond to the effect estimate of each IV to the exposure and outcome variable, respectively. The gradient of each regression line corresponds to the effect estimate obtained by each method (IVW, Egger, WM).

(top right) Forest plots of the effect estimates of each single IV on the outcome. Pooled effect estimate based on Egger and IVW methods are shown by the red bars.

(bottom left) Leave-one-out analysis. Red bar denotes the effect estimate by IVW.

(bottom right) Funnel plot used to gauge directional pleiotropy suggested by asymmetry.

All error bars correspond to 95% confidence intervals.

### Figure S12: Plots used to aid causal inference based on MR for sleep duration vs UPDRS3, without rs1633106

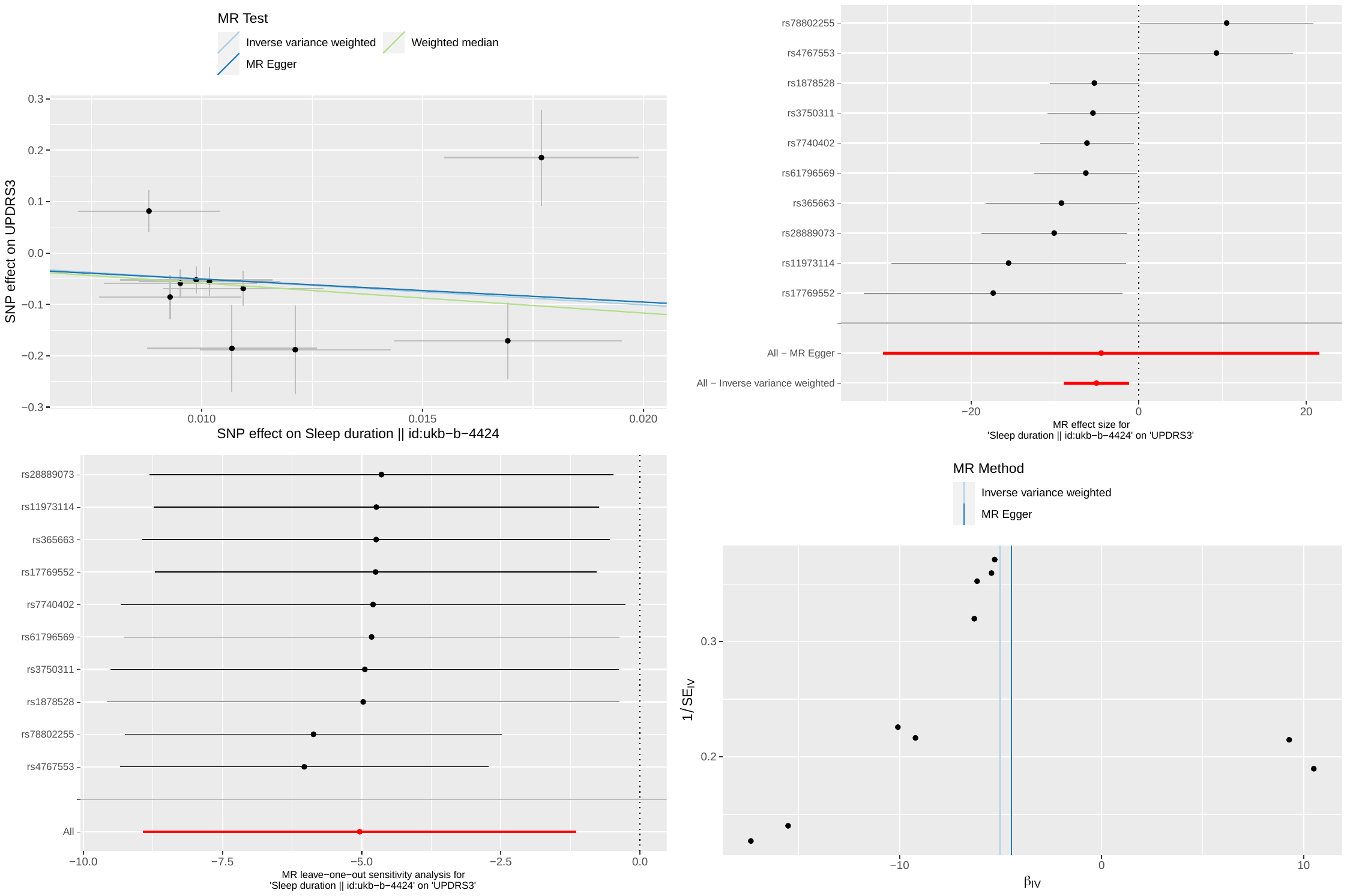

(top left) Scatter plot of the exposure (x-axis) and outcome (y-axis) variables of interest. Each point denotes a single IV included in the study. The x- and y-values of each point correspond to the effect estimate of each IV to the exposure and outcome variable, respectively. The gradient of each regression line corresponds to the effect estimate obtained by each method (IVW, Egger, WM).

(top right) Forest plots of the effect estimates of each single IV on the outcome. Pooled effect estimate based on Egger and IVW methods are shown by the red bars.

(bottom left) Leave-one-out analysis. Red bar denotes the effect estimate by IVW.

(bottom right) Funnel plot used to gauge directional pleiotropy suggested by asymmetry.

All error bars correspond to 95% confidence intervals.

### Figure S13: Plots used to aid causal inference based on MR for sleep duration vs MOCA

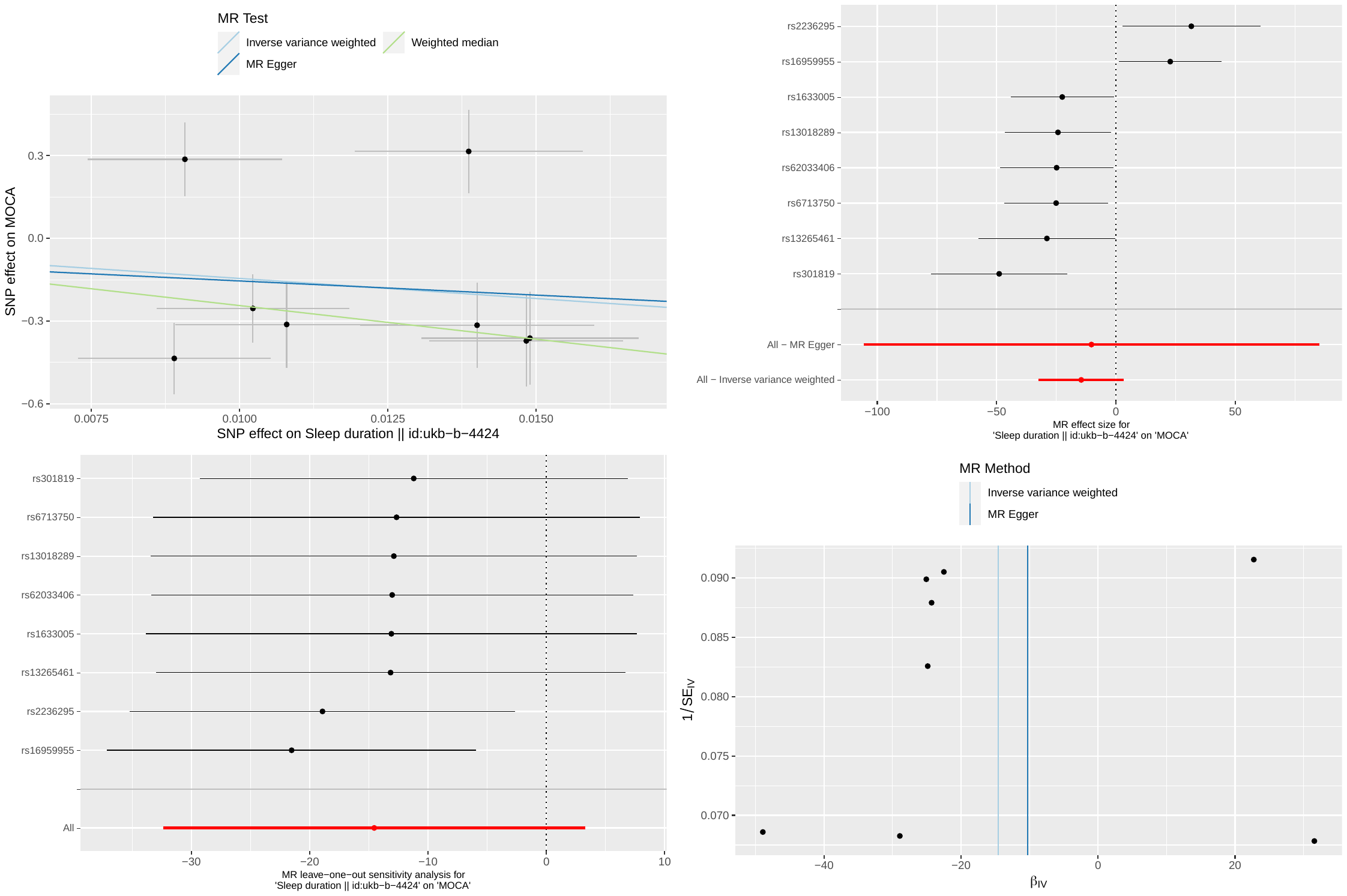

(top left) Scatter plot of the exposure (x-axis) and outcome (y-axis) variables of interest. Each point denotes a single IV included in the study. The x- and y-values of each point correspond to the effect estimate of each IV to the exposure and outcome variable, respectively. The gradient of each regression line corresponds to the effect estimate obtained by each method (IVW, Egger, WM).

(top right) Forest plots of the effect estimates of each single IV on the outcome. Pooled effect estimate based on Egger and IVW methods are shown by the red bars.

(bottom left) Leave-one-out analysis. Red bar denotes the effect estimate by IVW.

(bottom right) Funnel plot used to gauge directional pleiotropy suggested by asymmetry.

All error bars correspond to 95% confidence intervals.

### Figure S14: Plots used to aid causal inference based on MR for sleep duration vs MOCA, without rs2236295

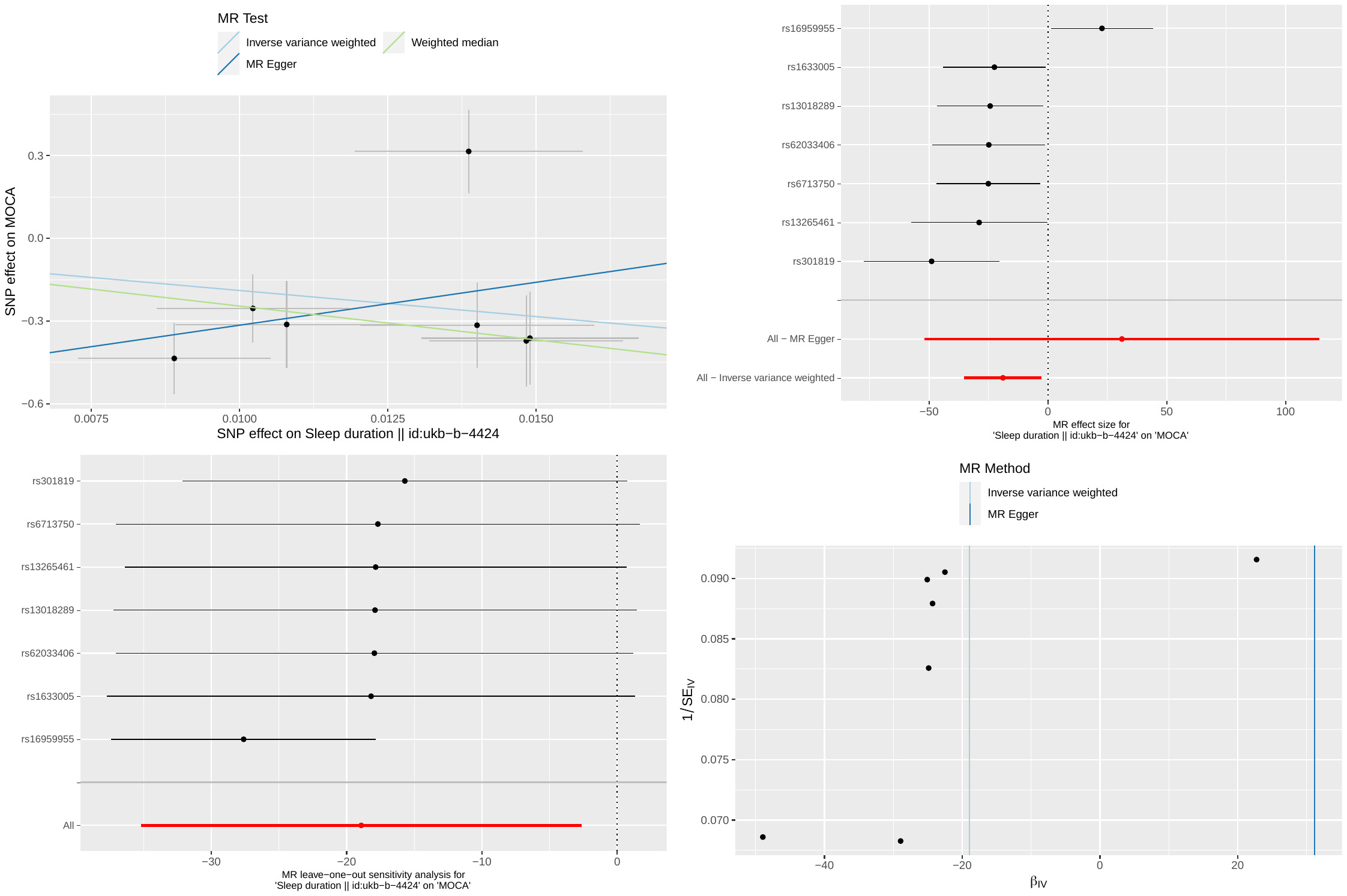

(top left) Scatter plot of the exposure (x-axis) and outcome (y-axis) variables of interest. Each point denotes a single IV included in the study. The x- and y-values of each point correspond to the effect estimate of each IV to the exposure and outcome variable, respectively. The gradient of each regression line corresponds to the effect estimate obtained by each method (IVW, Egger, WM).

(top right) Forest plots of the effect estimates of each single IV on the outcome. Pooled effect estimate based on Egger and IVW methods are shown by the red bars.

(bottom left) Leave-one-out analysis. Red bar denotes the effect estimate by IVW.

(bottom right) Funnel plot used to gauge directional pleiotropy suggested by asymmetry.

All error bars correspond to 95% confidence intervals.

### Figure S15: Plots used to aid causal inference based on MR for sleep duration vs MOCA, without rs16959955

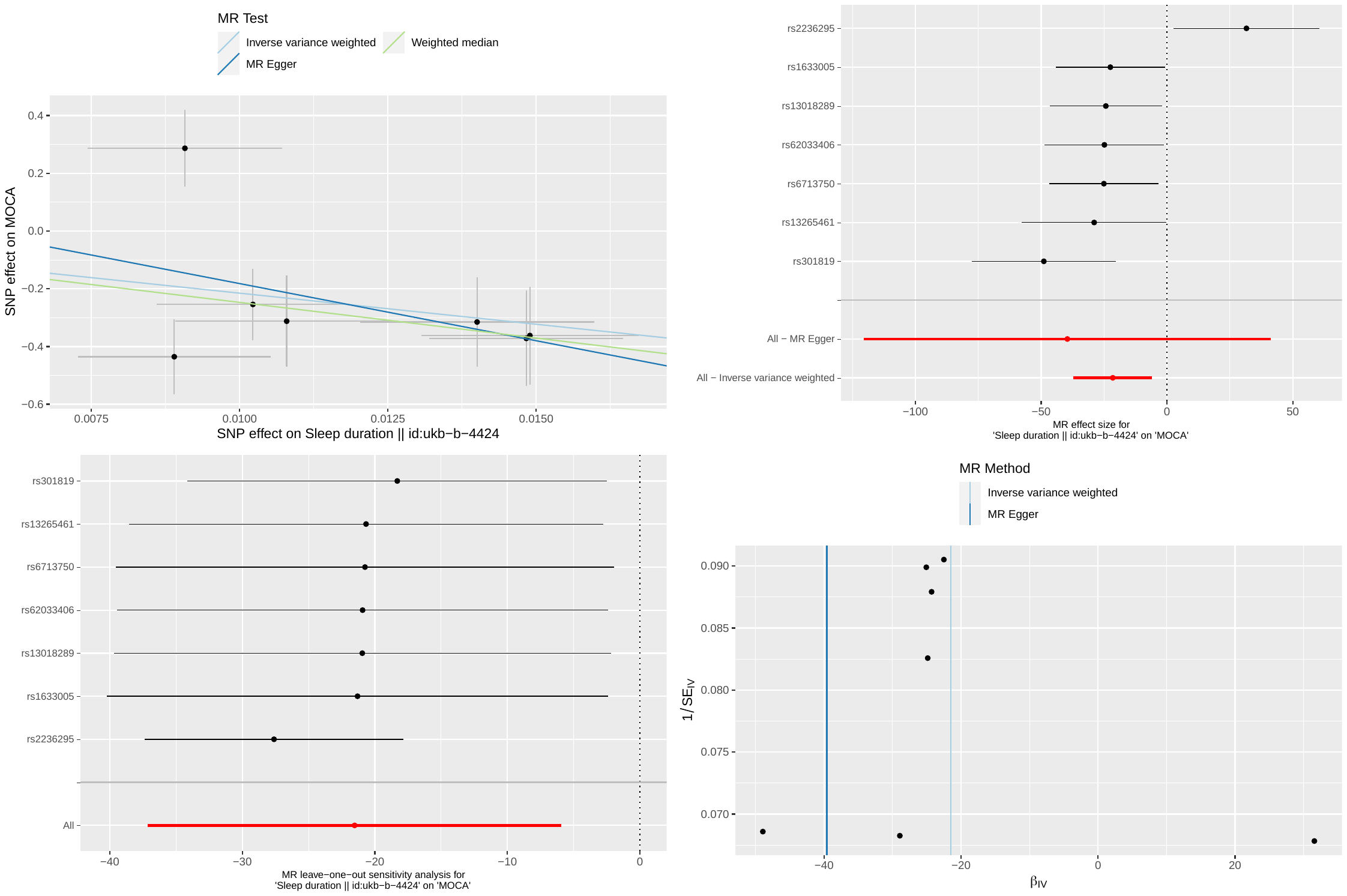

(top left) Scatter plot of the exposure (x-axis) and outcome (y-axis) variables of interest. Each point denotes a single IV included in the study. The x- and y-values of each point correspond to the effect estimate of each IV to the exposure and outcome variable, respectively. The gradient of each regression line corresponds to the effect estimate obtained by each method (IVW, Egger, WM).

(top right) Forest plots of the effect estimates of each single IV on the outcome. Pooled effect estimate based on Egger and IVW methods are shown by the red bars.

(bottom left) Leave-one-out analysis. Red bar denotes the effect estimate by IVW.

(bottom right) Funnel plot used to gauge directional pleiotropy suggested by asymmetry.

All error bars correspond to 95% confidence intervals.

### Figure S16: Plots used to aid causal inference based on MR for sleep duration vs MOCA, without rs16959955 and rs2236295

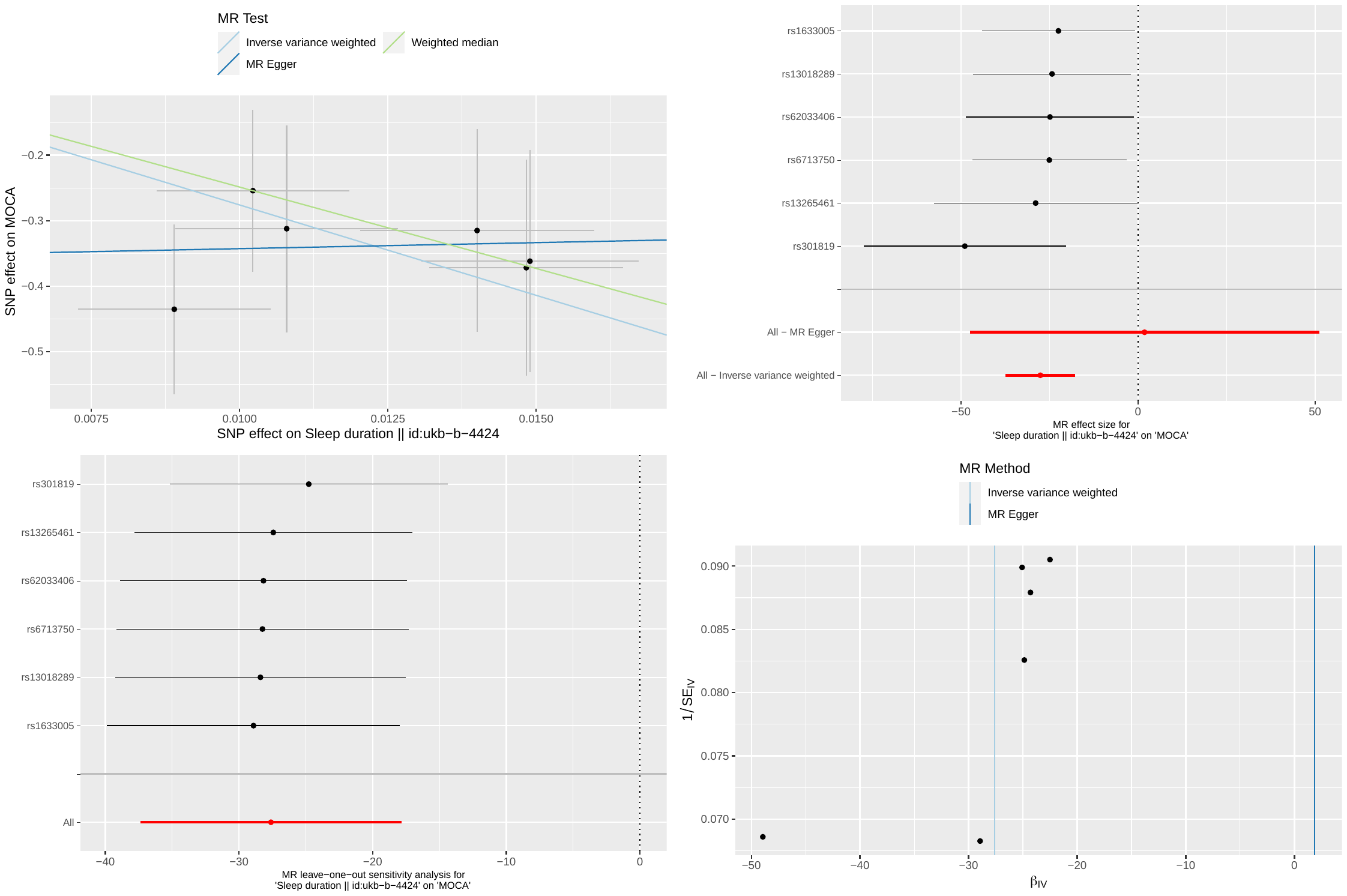

(top left) Scatter plot of the exposure (x-axis) and outcome (y-axis) variables of interest. Each point denotes a single IV included in the study. The x- and y-values of each point correspond to the effect estimate of each IV to the exposure and outcome variable, respectively. The gradient of each regression line corresponds to the effect estimate obtained by each method (IVW, Egger, WM).

(top right) Forest plots of the effect estimates of each single IV on the outcome. Pooled effect estimate based on Egger and IVW methods are shown by the red bars.

(bottom left) Leave-one-out analysis. Red bar denotes the effect estimate by IVW.

(bottom right) Funnel plot used to gauge directional pleiotropy suggested by asymmetry.

All error bars correspond to 95% confidence intervals.
